# Heme Oxygenase-1 Promoter (GT)n Alleles and Neurocognitive Functioning in Thai Youth with Perinatally-Acquired HIV: A Pilot Study

**DOI:** 10.1101/2022.04.22.22274113

**Authors:** Anthony F. Santoro, Linda Aurpibul, Dennis Kolson, Christopher M. Ferraris, Maral Aghvinian, Yoelvis Garcia-Mesa, Jun Lui, Sahera Dirajlal-Fargo, Reuben N. Robbins

## Abstract

**Background:** Heme oxygenase-1 (HO-1) gene promoter (GT)n dinucleotide repeat length variations may modify HIV-associated neurocognitive impairment (HIV-NCI) risk. Among adults, short HO-1 (GT)n alleles associate with greater HO-1 antioxidant enzyme inducibility and lower rates of HIV-NCI. This pilot study examined associations between HO-1 (GT)n alleles and neurocognitive outcomes in a sample of Thai youth (13-23 years) with perinatally-acquired HIV (PHIV) and demographically-matched HIV-negative controls.

**Methods:** Participants completed neurocognitive testing and provided blood samples for DNA extraction and sequencing of HO-1 promoter (GT)n dinucleotide repeat lengths. Allele lengths were assigned based on number of (GT)n repeats: <27 Short (S); 27-34 Medium (M); >34 Long (L). Relationships between HO-1 (GT)n repeat lengths and neurocognitive measures were examined, and differences by HO-1 (GT)n allele genotypes were explored.

**Results:** Nearly half (48%) of all HO-1 (GT)n promoter alleles were short. Longer repeat length of participants’ longest HO-1 (GT)n alleles significantly associated with poorer processing speed (Total sample: *r*=-.36, *p*=.01; PHIV only: *r*=-.69, *p*<.001). Compared to peers and controlling for covariates, SS/SM genotypes performed better in processing speed, and SS genotypes performed worse in working memory.

**Conclusions:** A high frequency of short HO-1 (GT)n alleles was found among these Thai youth, as previously observed in other cohorts of people of Asian ancestry. In contrast to previous adult studies, the presence of a short allele alone did not associate with better neurocognitive performance, suggesting additional modifying effects among the different alleles. Research is needed to determine whether HO-1 (GT)n promoter genotypes differentially influence neurocognitive functioning across the lifespan and different ethnic backgrounds.

---

HIV-associated neurocognitive impairment (HIV-NCI) is a common complication of HIV infection – particularly among youth living with perinatally-acquired HIV (PHIV)^1–3^. Upon infection, HIV crosses the blood-brain barrier, entering the central nervous system (CNS) primarily through transendothelial migration of HIV-infected CD4 T lymphocytes, with subsequent adaption to infect brain macrophages.^4,5^ In the CNS, HIV depletes immune cells and causes neural injury, leading to HIV-NCI.^5–12^ Although widespread access to antiretroviral therapy (ART) has reduced rates of classical HIV neuropathology (e.g., encephalitis), ART’s protection against HIV-NCI is incomplete.^5,13–15^ Remarkably, HIV-NCI still occurs in 35-59% of youth with PHIV despite long-term ART and well-controlled viremia.^16,17^ Among ART-treated youth, chronic neuroinflammation triggered by low-level viral replication and abnormal immune activation may drive ongoing neural injury.^6,12,18^ Further, in the context of viral suppression, non-HIV-specific host factors may play critical roles in HIV-NCI.^5,19–22^

Heme oxygenase-1 (HO-1) is a cytoprotective, detoxifying enzyme that may be a critical effector in reducing the CNS neuroinflammation, oxidative stress, and cellular injury observed in HIV-NCI.^21–30^ HO-1 is rapidly induced in response to cellular injury in the CNS and functions to degrade heme, generating carbon monoxide, biliverdin, and bilirubin.^31^ In turn, these processes have anti-oxidative effects and help modulate immune activation,^31^ protecting the CNS from further injury.^32,33^ Among adults with HIV, decreased HO-1 protein expression associates with increased immune activation in the CNS, including interferon responses (i.e., interferon gamma-soluble cytokine [IFN-γ]).^21^ Furthermore, reduced HO-1 expression in HIV-infected macrophages may augment neurotoxins produced by these macrophages,^21,23^ suggesting reduced HO-1 expression may promote CNS injury via heightened neurotoxin production.^22,23^ Collectively, evidence suggests important relationships between reduced HO-1 in the brain, immune activation, and HIV-associated CNS dysfunction.^22^ Notably, HO-1 expression can be induced pharmacologically^28,29^ – allowing for exciting new possibilities for developing biomedical therapies for treating, and perhaps preventing, HIV-NCI.^29^

HO-1 expression is regulated by genetic polymorphisms in the human HO-1 gene (*HMOX1*).^22,34^ One such polymorphism, the HO-1 (GT)n dinucleotide repeat length in the *HMOX1* promotor region (i.e., 5′-flanking region), has been shown to modulate HO-1 promoter activity.^35–37^ Specifically, shorter (GT)n repeat lengths associate with higher HO-1 expression in response to oxidative stress^35–39^ and better clinical outcomes in inflammation- and oxidative stress-related diseases, including coronary artery disease,^35^ ischemic heart disease and stroke,^37,38^ and rheumatoid arthritis.^39^ Among adults with HIV, shorter HO-1 (GT)n repeat lengths have been associated with lower viral load and soluble CD14,^24^ a key marker of immune activation associated with HIV-NCI.^40^ Autopsy studies suggest shorter HO-1 (GT)n alleles associate with lower rates of HIV encephalitis and lower neuroinflammation markers (i.e., type I interferon response, T-cell activation).^22^ Moreover, in a large well-characterized cohort of adults living with HIV, short HO-1 (GT)n allele genotypes were found to associate with lower rates of HIV-NCI.^41^ Taken together, evidence suggests HO-1 (GT)n allele repeat length may serve as a unique genetic-level modifier of HIV-NCI.^20–22^

However, previous studies have largely focused on middle-aged adults with HIV in the United States, and studies among younger samples are needed to determine whether associations between HO-1 (GT)n polymorphisms and HIV-NCI are consistent across the lifespan or emerge during distinct developmental periods. Such information is critical to informing the timing of future prevention efforts for youth who may exhibit genetic vulnerabilities for developing HIV-NCI. Likewise, HO-1 (GT)n allele repeat lengths vary significantly across ethnicities, suggesting genetic vulnerabilities for HIV-NCI may vary across populations, and there is need for further research among additional ethnically distinct samples of people living with HIV.^41^ For example, despite about 10% of the world-wide HIV population living in Asian/Pacific resource-limited countries,^42^ the HO-1 (GT)n polymorphism remains understudied among populations of Asian ancestry, limiting our understanding of relative genetic risk of HIV-NCI within distinct Asian ethnic groups. That said, studies of HIV-negative adults of Asian ancestry have linked the presence of longer HO-1 (GT)n repeats with increased risk for chronic emphysema in smokers,^43^ and the presence of short HO-1 (GT)n alleles with decreased risk for coronary heart disease^35,44^ and increased susceptibility to cerebral malaria.^45^

To date, no published studies have addressed the role of HO-1 (GT)n promoter alleles in clinical outcomes in HIV populations of Asian ancestry, and associations between HO-1 (GT)n polymorphisms and neurocognitive outcomes have yet to be examined among young people living with PHIV. Considering these research gaps, this pilot study aimed to (1) provide preliminary estimates of HO-1 (GT)n allele lengths and genotypes and (2) explore associations between HO-1 (GT)n allele lengths and neurocognitive outcomes in a sample of Thai adolescents and young adults with PHIV and demographically matched HIV-negative peers. Based on previous studies, we expected that shorter HO-1 (GT)n alleles and genotypes would associate with better neurocognitive outcomes.

## Methods

### Design, Setting, and Participants

This pilot study recruited fifty Thai adolescents and young adults (13-23 years) with PHIV and age- and sex-matched HIV-negative controls from an ongoing parent study at the Research Institute for Health Sciences, Chiang Mai University (RIHES-CMU) in Thailand. As part of the parent study, participants completed a battery of tablet-based neurocognitive tests. Parent study eligibility included: a) 13-23 years old; b) PHIV; c) currently on an ART regimen. PHIV participants who previously completed the parent study were recruited to participate in this pilot study during regular visits at the ART clinic. Demographically matched (i.e., age, sex) HIV-negative control participants were recruited from communities in Chiang Mai city. One control participant declined genetic testing, yielding a final sample of 49 participants (25 PHIV; 24 controls).

### Procedures

This study was approved by the New York State Psychiatric Institute’s IRB and RIHES-CMU’s Human Experimentation Committee. Written informed consent was collected from participants ≥18 years. For participants <18 years, participant assent and written informed consent from a parent/guardian were collected. Following informed consent, participants completed a blood draw, collecting 2ml of whole blood via venipuncture. Blood samples were stored in a -80C freezer and cold-chain shipped in one batch for genetic testing. All study materials (e.g., consent forms) were translated into Thai. Demographic, viral load, CD4 count, and neurocognitive testing data were provided by the parent study.

### Measures

#### HO-1 (GT)n Promoter Genotype

Genomic DNA was extracted from whole blood samples (DNA Extraction Kit, Agilent Technologies), following previously described protocols for amplification and determination of genotype.^22,41^ Briefly, HO-1 (GT)n repeat lengths were determined by polymerase chain reaction (PCR) amplification of the (GT)n repeat region with a 6-fluorescein amidites (FAM) 5’ labeled forward primer (5′-FAM-CCAGCTTTCTGGAACCTTCTG-3′), and unlabeled reverse primer (5′-GAAACAAAGTCTGGCCATAGGA-3′), followed by fragment size determination on a capillary sequencer. For a subgroup of samples, PCR products were run on a 2% agarose gel to assess amplification of the target sequence predicted sizes (homozygotes and heterozygotes). Each sample was run at least twice from independent PCR reactions to confirm accurate and reproducible sizing, which was determined to be accurate if within ±1 GT repeats. If identical (GT)n repeat lengths were not rendered in duplicate, samples were run additional times to confirm repeat lengths. Alleles were classified by the number of (GT)n repeats: <27 (Short [S]); 27-34 (Medium [M]); >34 (Long [L]). Based on length of each participant’s two HO-1 (GT)n alleles, participants were assigned genotypes (i.e., SS, SM, SL, MM, ML, LL). For example, the SS genotype was assigned to participants with two short alleles.

#### Neurocognitive Measures

As part of the parent study, participants completed *NeuroScreen*, a neuropsychological battery of tablet-based tests measuring processing speed, working memory, and motor dexterity. *NeuroScreen* has been culturally- and linguistically-adapted for use among Thai-speaking adolescents and young adults.^46^ This pilot analyzed data from 10 *NeuroScreen* tests: five processing speed tests (Trail Making 1, Trail Making 3, Visual Discrimination 1, Visual Discrimination 2, Number Speed), one executive functioning test (Trail Making 2), two working memory tests (Number Span Forward, Number Span Backward), and two motor tests (Finger Tapping Dominant Hand, Finger Tapping Nondominant Hand). Detailed descriptions of *NeuroScreen* tests have been previously reported.^47^ Raw test scores were converted to demographically-adjusted (i.e., age, sex, education) T scores (*M* = 50, *SD* = 10).

### Analytic Plan

Descriptive statistics were generated to summarize demographic and genotype data. Histograms and bar charts were produced to visualize the frequency distributions of HO-1 (GT)n repeat lengths and allele genotypes. Spearman’s rho correlations were assessed associations between HO-1 (GT)n allele repeat lengths as continuous variables and neurocognitive outcomes for the total sample and PHIV participants only, bootstrapping 5,000 samples and calculating 95% confidence intervals (95% CI). As exploratory analyses, preliminary nonparametric Mann-Whitney *U* tests and general linear models (GLM), controlling for age, sex, education, and HIV status, explored differences between HO-1 (GT)n allele genotypes on neurocognitive outcomes. Considering Mann-Whitney *U* tests and GLMs yielded similar results, GLM results are reported. In preliminary analyses, no genotype by HIV status interactions were found; thus, the full sample was used to preserve sample size, entering HIV status as a covariate. Parameter estimates with robust standard errors (HC4 method) were produced for planned contrasts of interest, comparing: a) participants with and without at least one short allele (SS/SM/SL vs. MM/ML), b) the two shortest genotype groups and all other genotypes (SS/SM vs. SL/MM/ML), c) participants with two short alleles and all other genotypes (SS vs. SM/SL/MM/ML). Effect sizes were calculated 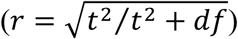.^48^ For completeness, GLMs were also run restricting analyses to PHIV participants only.

## Results

### Participant Characteristics

Participants’ demographics are summarized in Table 1. The average age of participants was 18.29 years old; 53.1% were female, and 83.7% were right-handed. PHIV and control participants did not differ regarding age, biological sex, or handedness. Years of education were higher among control participants (*M* = 12.21) than PHIV participants (*M* = 10.56). On average, PHIV participants have 13.90 years of ART duration, with 95% having ≥9 years of ART. Regarding PHIV participants’ most recent viral loads and CD4 counts, 79.2% had viral loads <200 copies/mL and 72% had CD4 counts >500 cells/mm^3^.

**Table 1.** Demographic Characteristics of Study Participants (N=49)

|  | **PHIV (n=25)** | **Controls (n=24)** | **Total (N=49)** |
| --- | --- | --- | --- |
|  | **% (n)** | **% (n)** | **% (n)** |
| **Age (years)** | *M*=18.28 (*SD*=3.09) | *M*=18.29 (*SD*=3.09) | *M*=18.29 (*SD*=3.06) |
| 13-17 years | 44.0% (11) | 45.8% (11) | 44.9% (22) |
| 18-23 years | 56.0% (14) | 54.2% (13) | 55.1% (27) |
| **Biological Sex** |  |  |  |
| Female | 52.0% (13) | 54.2% (13) | 53.1% (26) |
| Male | 48.0% (12) | 45.8% (11) | 46.9% (23) |
| **Education (years)** | *M*=10.56 (*SD*=2.74) | *M*=12.21 (*SD*=2.92) | *M*=11.37 (*SD*=2.92) |
| ≤9 years | 36.0% (9) | 20.8% (5) | 28.6% (14) |
| 10-12 years | 24.0% (6) | 12.5% (3) | 18.4% (9) |
| ≥12 years | 40.0% (10) | 66.7% (16) | 53.1% (26) |
| **Handedness** |  |  |  |
| Right | 84.0% (21) | 83.3% (20) | 83.7% (41) |
| Left | 16.0% (4) | 16.7% (4) | 16.3% (8) |
| **ART Duration (years)<sup>c</sup>** | *M*=13.90 (*SD*=2.75) | — | — |
| **Current ART Regimen<sup>c</sup>** |  |  |  |
| EFV+TDF+FTC | 57.1% (12) | — | — |
| Other Regimen | 42.9% (9) | — | — |
| **Recent Viral Load<sup>a</sup>** |  |  |  |
| <200 copies/mL | 79.2% (19) | — | — |
| >200 copies/mL | 20.8% (5) | — | — |
| **Recent CD4 Count<sup>b</sup>** |  |  |  |
| <200 cells/mm<sup>3</sup> | 8.0% (2) | — | — |
| 200-500 cells/mm<sup>3</sup> | 20.0% (5) | — | — |
| >500 cells/mm<sup>3</sup> | 72.0% (18) | — | — |

Participant demographic data was provided by the parent study.
a. Recent viral load data was available for 24 PHIV participants. On average, there were 190 days between most recent viral load and neurocognitive testing.
b. Recent CD4 count data was available for 25 PHIV participants. On average, there were 201 days between most recent CD4 count and neurocognitive testing.
c. ART start date and current ART regimen data was available for 21 PHIV participants.
EFV+TDF+FTC = Efavirenz + Tenofovir + Emtricitabine

### HO-1 (GT)n Allele Length and Genotypes

Figure 1 depicts the distribution and prevalence of HO-1 (GT)n promoter allele lengths in the current sample. A high number of short alleles was found, with nearly half (48.0%) of all HO-1 (GT)n promoter allele lengths characterized as short. Conversely, few long alleles (8.2%) were found in the current sample.

**Figure 1.**
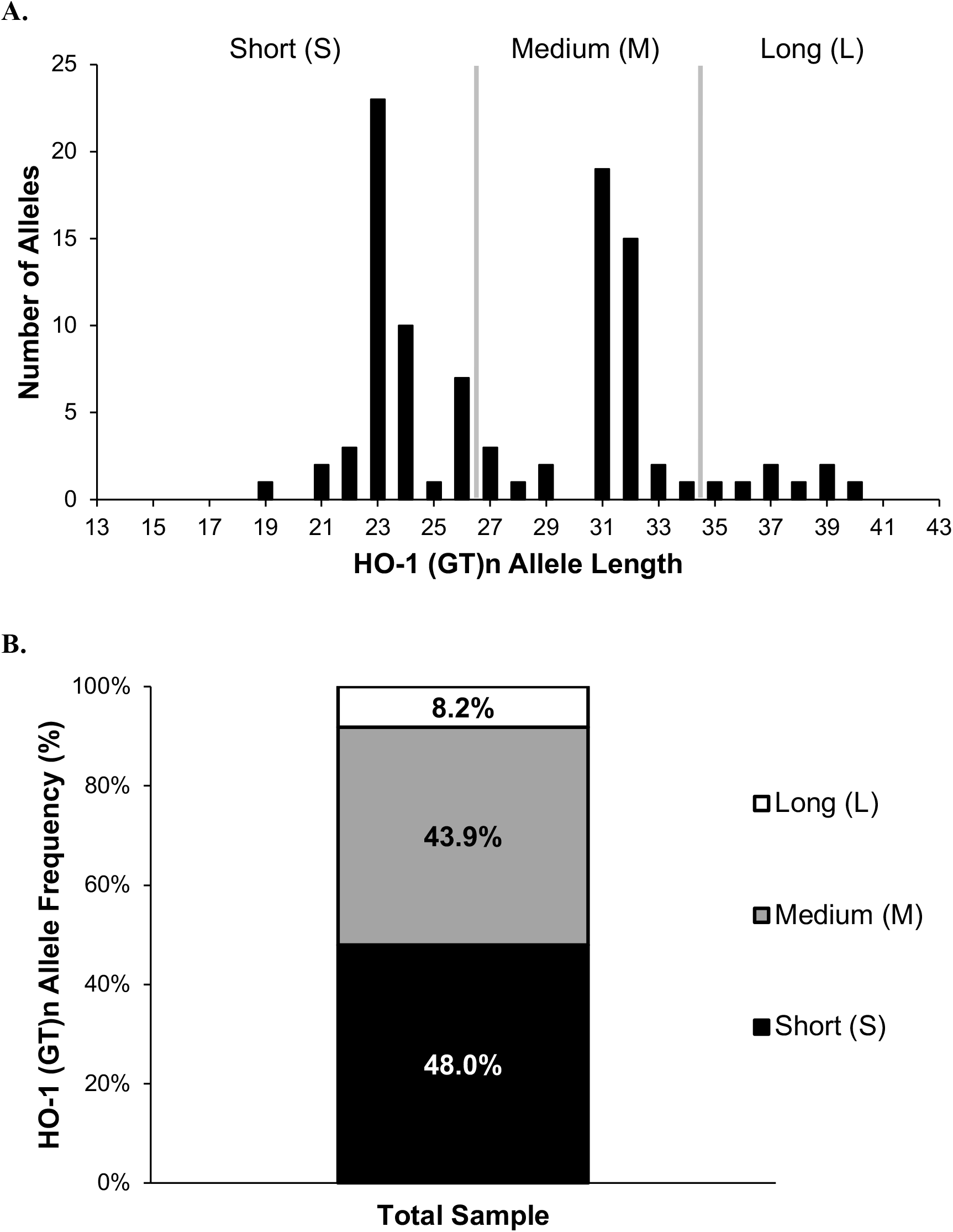
Distribution (A) and frequency (B) of HO-1 (GT)n alleles. Allele lengths were classified based on (GT)n repeats as short “S” (<27 repeats), medium “M” (27–34 repeats), and long “L” (>34 repeats).

Figure 2 depicts the frequency of HO-1 (GT)n genotypes. Nearly three-fourths (73.5%) of participants had at least one short allele (SS/SM/SL), and 22.4% had two short alleles (SS genotype). Only 16.3% of participants had a long allele (SL/ML), and no participants had two long alleles (LL). Regarding genotype prevalence, SM (42.9%) and SS (22.4%) genotypes were most prevalent. Among genotypes present in the sample, SL (8.2%) and ML (8.2%) genotypes were the least common. Similar prevalence of HO-1 (GT)n genotypes was found between the PHIV and control groups.

**Figure 2.**
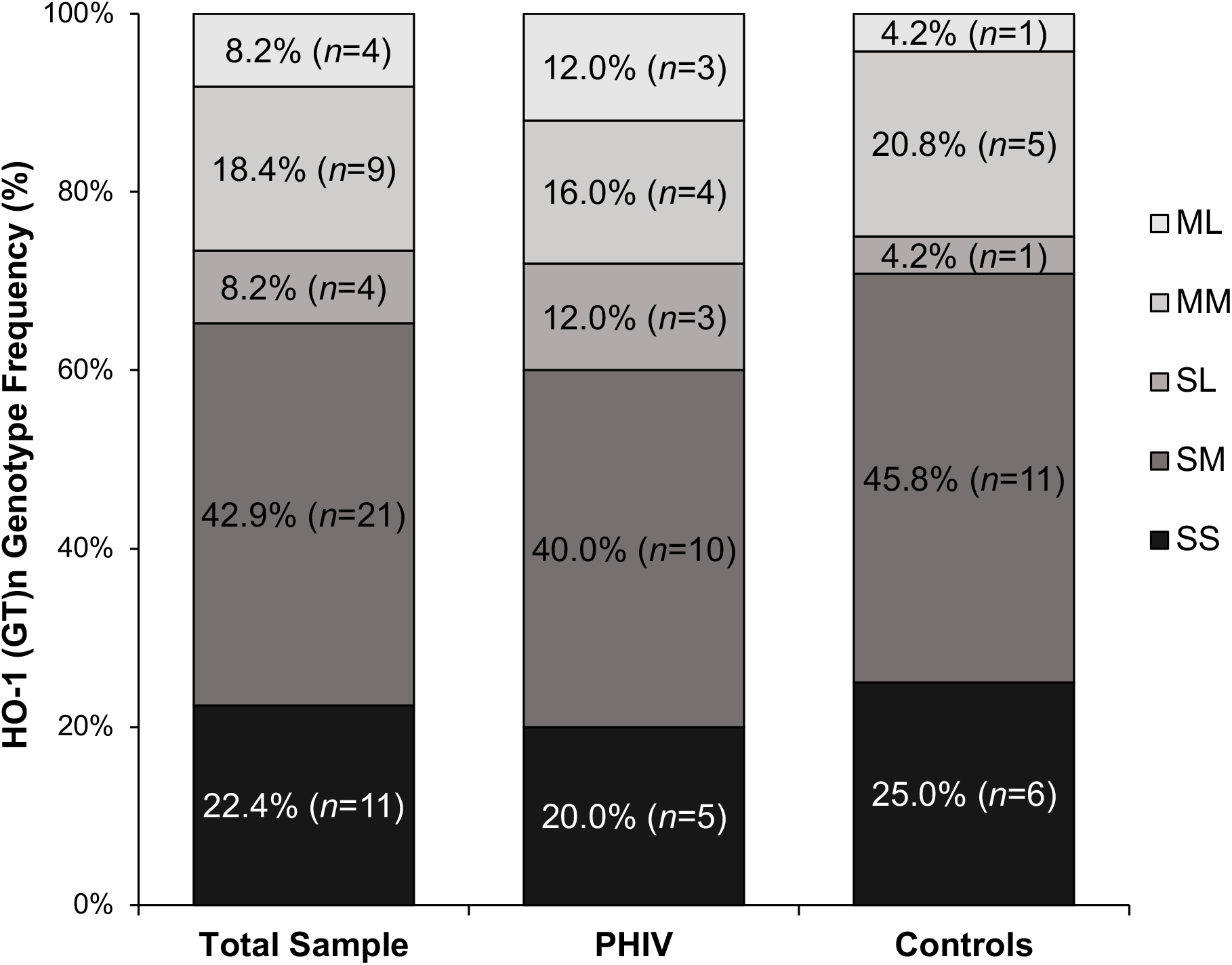
Frequency of HO-1 (GT)n genotypes. The SM genotype was most prevalent. No participant had two long alleles (i.e., LL genotype).

### Correlations between HO-1 (GT)n Allele Length and Neurocognitive Measures

Analyzing HO-1 (GT)n allele lengths as continuous variables, in the total sample, longer repeat lengths of participants’ longest allele (i.e., number of repeats in participants’ least expressive HO-1 promoter) was significantly associated with worse performance on Trail Making 3 (*r* = -.36, *p* = .01), a measure of simple processing speed. Among the total sample, longest allele length also trended toward significance for Trail Making 1 (*p* = .06); no significant associations were found between shortest allele length and neurocognitive measures. Restricting analyzes to PHIV participants, longer repeat lengths of participants’ longest allele was also significantly associated with worse performance on Trail Making 3 (*r* = -.69, *p* < .001). Additionally, among PHIV participants, the association between shortest allele length and Trail Making 3 approached significance (*p* = .054), with repeat length inversely related to processing speed performance. Correlation coefficients with 95% CIs are presented in Table 2.

**Table 2.** Correlation Coefficients between Shortest and Longest HO-1 (GT)n Allele Lengths and Neurocognitive Outcomes.

|  |  | Shortest Allele Length |  |  | Longest Allele Length |  |  |
| --- | --- | --- | --- | --- | --- | --- | --- |
|  |  | *r* | [95% CI] | *p* | *r* | [95% CI] | *p* |
| TM 1 | Total: | -.04 | [-.32, .24] | *ns* | -.27 | [-.53, .03] | .06 |
|  | PHIV Only: | .22 | [-.26, .62] | *ns* | -.22 | [-.66, .30] | *ns* |
| TM 2 | Total: | .12 | [-.18, .41] | *ns* | .04 | [-.24, .32] | *ns* |
|  | PHIV Only: | .26 | [-.19, .64] | *ns* | .03 | [-.41, .44] | *ns* |
| TM 3 | Total: | -.16 | [-.45, .14] | *ns* | -.36 | [-.61, -.08] | **<u>.01</u>** |
|  | PHIV Only: | -.39 | [-.75, .07] | .054 | -.69 | [-.88, -.40] | **<u><.001</u>** |
| VD 1 | Total: | .15 | [-.11, .40] | *ns* | .10 | [-.18, .36] | *ns* |
|  | PHIV Only: | .11 | [-.27, .49] | *ns* | .15 | [-.23, .50] | *ns* |
| VD 2 | Total: | -.05 | [-.31, .23] | *ns* | -.08 | [-.36, .22] | *ns* |
|  | PHIV Only: | -.10 | [-.49, .31] | *ns* | -.10 | [-.53, .35] | *ns* |
| NSPD | Total: | -.13 | [-.45, .20] | *ns* | -.17 | [-.43, .12] | *ns* |
|  | PHIV Only: | .19 | [-.24, .56] | *ns* | -.02 | [-.41, .36] | *ns* |
| NSPN F | Total: | .04 | [-.27, .33] | *ns* | .25 | [-.04, .52] | .08 |
|  | PHIV Only: | .18 | [-.30, .57] | *ns* | .26 | [-.21, .67] | *ns* |
| NSPN B | Total: | .06 | [-.25, .35] | *ns* | .02 | [-.27, .31] | *ns* |
|  | PHIV Only: | -.11 | [-.53, .32] | *ns* | .04 | [-.38, .44] | *ns* |
| FT D | Total: | -.11 | [-.38, .17] | *ns* | .01 | [-.27, .31] | *ns* |
|  | PHIV Only: | .12 | [-.31, .57] | *ns* | .04 | [-.39, .49] | *ns* |
| FT ND | Total: | -.12 | [-.40, .17] | *ns* | -.10 | [-.37, .19] | *ns* |
|  | PHIV Only: | .05 | [-.40, .51] | *ns* | -.13 | [-.50, .32] | *ns* |

*Note.* Spearman rho correlation coefficients were conducted bootstrapping 5,000 samples for the total sample and for PHIV participants only (shaded grey). Significant *p*-values ( $p < .05$ ) are bolded and underlined. 95% CI = 95% confidence intervals; *ns* = non-significant *p*-values $\geq .10$ . **Processing speed:** TM 1 = Trail Making 1; TM 3 = Trail Making 3; VD 1 = Visual Discrimination 1; VD 2 = Visual Discrimination 2; NSPD = Number Speed. **Executive functioning:** TM 2 = Trail Making 2. **Working memory:** NSPN F = Number Span Forward; NSPN B = Number Span Backward. **Motor dexterity:** FT D = Finger Tapping Dominant Hand; FT ND = Finger Tapping Nondominant Hand.

### Comparing Neurocognitive Outcomes by HO-1 (GT)n Genotype Groups

Table 3 presents parameter estimates exploring differences between HO-1 (GT)n genotype groupings. Controlling for age, sex, education, and HIV status, membership in the shortest genotype groups (SS/SM) had a significant effect on Trail Making 3 performance, *F*(1, 43) = 5.12, *p* = .03, and this effect approached significance for Number Span Forward, *F*(1, 43) = 3.91, *p* = .054. On Trail Making 3, planned contrast revealed SS/SM genotypes (*M* = 48.25, *SD* = 8.51) exhibited better simple processing speed than SL/MM/ML genotypes (*M* = 41.82, *SD* = 9.13), *t*(43) = -2.19, *p* = .03, *r* = .32. Conversely, on Number Span Forward, participants with SS/SM genotypes (*M* = 43.09, *SD* = 11.10) exhibited worse working memory than SL/MM/ML genotypes (*M* = 47.94, *SD* = 8.27), *t*(43) = 2.15, *p* = .04, *r* = .31. Separately, the presence of two short alleles (SS) has a significant effect on Number Span Forward performance, *F*(1, 43) = 11.66, *p* = .001, with SS genotypes (*M* = 38.09, *SD* = 11.87) exhibiting worse working memory than other genotypes (*M* = 46.71, *SD* = 9.20), *t*(43) = 2.87, *p* = .006, *r* = .40.

**Table 3.** Differences in Neurocognitive Measures by HO-1 (GT)n Genotypes.

|  |  | SS/SM vs. SL/ML/MM |  |  | SS vs. SM/SL/MM/ML |  |  |
| --- | --- | --- | --- | --- | --- | --- | --- |
|  |  | *t* | *p* | *r* (*ES*) | *t* | *p* | *r* (*ES*) |
| TM 1 | Total: | -1.08 | *ns* | — | 0.14 | *ns* | — |
|  | PHIV Only: | -0.21 | *ns* | — | -0.86 | *ns* | — |
| TM 2 | Total: | 0.42 | *ns* | — | 1.35 | *ns* | — |
|  | PHIV Only: | 1.42 | *ns* | — | 0.32 | *ns* | — |
| TM 3 | Total: | -2.19 | **.03** | .32 (M) | -1.02 | *ns* | — |
|  | PHIV Only: | -3.45 | **.003** | .61 (L) | -2.17 | **.04** | .44 (M) |
| VD 1 | Total: | 1.74 | .09 | .26 (S/M) | 1.85 | .07 | .27 (S/M) |
|  | PHIV Only: | 1.35 | *ns* | — | 0.85 | *ns* | — |
| VD 2 | Total: | -0.21 | *ns* | — | 0.96 | *ns* | — |
|  | PHIV Only: | -0.50 | *ns* | — | 0.28 | *ns* | — |
| NSPD | Total: | -0.18 | *ns* | — | -0.78 | *ns* | — |
|  | PHIV Only: | 0.30 | *ns* | — | 0.58 | *ns* | — |
| NSPN F | Total: | 2.15 | **.04** | .31 (M) | 2.87 | **.006** | .40 (M) |
|  | PHIV Only: | 1.01 | *ns* | — | 0.74 | *ns* | — |
| NSPN B | Total: | 0.49 | *ns* | — | 1.75 | .09 | .26 (S/M) |
|  | PHIV Only: | -0.09 | *ns* | — | 2.02 | .06 | .42 (M) |
| FT D | Total: | -0.94 | *ns* | — | 0.87 | *ns* | — |
|  | PHIV Only: | -0.13 | *ns* | — | 0.73 | *ns* | — |
| FT ND | Total: | -0.89 | *ns* | — | 1.30 | *ns* | — |
|  | PHIV Only: | -0.44 | *ns* | — | 0.81 | *ns* | — |

*Note.* Differences by genotype groups for the total sample and PHIV participants only (shaded gray). General linear models entered age, sex, education, and HIV status as covariates.
Significant *p*-values ( $p < .05$ ) are bolded. Effect sizes (*r*) are presented for results with $p < .10$ .
Effect sizes of .10, .30, and .50 were considered small, medium, and large, respectively.<sup>50</sup>
*ES* = effect size; *ns* = non-significant *p*-values $\geq .10$
Processing speed: TM 1 = Trail Making 1; TM 3 = Trail Making 3; VD 1 = Visual
Discrimination 1; VD 2 = Visual Discrimination 2; NSPD = Number Speed.
Executive functioning: TM 2 = Trail Making 2. Working memory: NSPN F = Number Span
Forward; NSPN B = Number Span Backward. Motor dexterity: FT D = Finger Tapping
Dominant Hand; FT ND = Finger Tapping Nondominant Hand.

Restricting analyses to PHIV participants, membership in the shortest genotype groups (SS/SM) was also significantly associated with Trail Making 3 performance, *F*(1, 20) = 10.39, *p* = .004; PHIV participants with SS/SM genotypes (*M* = 47.40, *SD* = 6.56) exhibited better simple processing speed than SL/MM/ML genotypes (*M* = 39.10, *SD* = 5.74), *t*(20) = -3.45, *p* = .003, *r* = .61, representing a large effect size. Separately, the presence of two short alleles was also associated with Trail Making 3 performance, *F*(1, 20) = 6.58, *p* = .02; PHIV participants with SS genotypes (*M* = 51.40, *SD* = 7.16) exhibited better simple processing speed than other genotypes (*M* = 42.25, *SD* = 6.37), *t*(20) = -2.17, *p* = .04, *r* = .44. The presence/absence of at least one short allele (SS/SM/SL vs. MM/ML) was not significantly associated with neurocognitive measures in models using either the total sample or restricted to PHIV participants.

## Discussion

In this Thai adolescent and young adult sample, the prevalence and distributions of HO-1 (GT)n promoter alleles and genotypes were distinct compared to those from White American and African American samples of adults with HIV.^41^ In our sample, most participants had at least one short allele, and no participant had two long alleles. This high frequency of short alleles is similar to previous reports in HIV-negative adults from Myanmar, bordering Thailand.^45^ These data add to the evidence that HO-1 allele lengths may vary significantly across geographic region, highlighting the need for further genetic studies with ethnically and regionally diverse populations of young people with HIV. Moreover, data suggest genetic risk of HIV-NCI may vary across distinct ethnic groups, with populations with a greater prevalence of longer alleles potentially carrying relatively higher risk.

Associations between HO-1 (GT)n genotypes and neurocognitive outcomes were also distinct. Even in this small sample, we found significant associations between HO-1 (GT)n allele lengths and neurocognitive outcomes. Specifically, longer repeat lengths of participants’ longest (GT)n allele (i.e., number of repeats of participant’s least expressive/inductive HO-1 promoter) associated with worse simple processing speed. This association represented a large effect size when analyses were restricted to PHIV participants. Additionally, associations between allele length and a separate measure of processing speed and a measure of working memory approached significance, suggesting further investigation of these relationships in larger sample size is warranted.

Previous studies of adults with HIV have found that the presence of a short HO-1 (GT)n allele associates with a lower prevalence of HIV-NCI;^41^ however, in our sample, the presence of a short allele alone was not significantly associated with neurocognitive outcomes. That said, short HO-1 genotypes SS and SM performed better in simple processing speed (i.e., Trail Making 3) than other genotypes, and this difference represented a large effect size among PHIV participants. Notably, grouping participants by presence/absence of a short allele sorted those with both a short and long allele (i.e., SL) in the “short” group, and associations with cognitive outcomes failed to reach significance. However, when the “short” group was restricted to the two shortest genotypes (i.e., SS and SM), excluding SL genotypes, we found membership in the “short” group significantly associated with better processing speed. Taken together, findings suggest that the presence of a short allele benefited participants’ processing speed if also in the absence of a long allele.

Unexpectedly, SS genotypes performed worse than peers in working memory. Considering these participants have two short HO-1 (GT)n alleles, which associates with greatest HO-1 expression,^35–39^ we would expect these participants are afforded the greatest neuroprotection. Thus, we expected SS genotypes to associate with better neurocognitive functions. Intriguingly, we found the opposite in our total sample, with SS genotypes performing worse than peers with genotypes that less readily express HO-1. In contrast, a study of 276 adults of African ancestry living with HIV showed that those with at least one short allele had a significantly reduced risk of HIV-NCI (defined across seven neurocognitive domains).^41^ Discrepancies between current and previous findings could be related to the current sample including both PHIV and HIV-negative participants, whereas previous adult studies have not included HIV-negative controls. That said, discrepancies could also be related to variations across the lifespan and between ethnic groups. Although the scarcity of literature precludes robust comparisons, one study of 28 Ugandan adolescents with PHIV found SS genotypes associated with worse executive functioning and complex processing speed.^49^ Thus, additional studies are needed to determine how HO-1 (GT)n promoter genotypes may differentially influence neurocognitive functioning across the lifespan and within different ethnic backgrounds.

This study has several limitations. This was a pilot study with a small sample size, which precluded examining differences across all genotype groups. Additionally, analyses were exploratory, and analyses restricted to only PHIV participants were likely insufficiently powered. Considering that we found several associations that approached significance, additional associations between genotypes and specific neurocognitive outcomes may exist in this population that exploratory analyses were not adequately powered to detect. Studies with larger sample sizes are needed to corroborate these preliminary findings and examine additional comparisons of interest.

Despite limitations, this pilot study significantly contributes to the literature. To our knowledge, it is among the first to examine the prevalence of HO-1 (GT)n alleles and associations with neurocognitive functions in a Thai sample. It is also among the first to explore these relationships in adolescents and young adults living with PHIV. Likewise, a strength of this pilot was its use of standardized neurocognitive measures, extending the literature in identifying specific neurocognitive domains that may be particularly sensitive to variations in HO-1 (GT)n alleles, as well as a comparator group that was demographically similar and from the same region. These preliminary findings are hoped to inform new and exciting veins of research that have the potential to drive advancements in treating HIV-NCI, specifically among youth living with PHIV in resource-limited settings. Additional larger studies examining HO-1 (GT)n genotype distributions among other distinct Asian populations of young people living with HIV are warranted.

## Data Availability

All data produced in the present study are available upon reasonable request to the authors.

## Acknowledgments

We would like to acknowledge all our participants and the research team at the Research Institute for Health Sciences, Chiang Mai University.

## Authors’ Contributions

AFS led the study conceptualization and design, data analyzes, writing the original draft, and revising the manuscript. LA oversaw data collection, supervised the study team in Thailand, and reviewed and revised the manuscript draft. DK provided mentorship and reviewed and revised the manuscript draft. CMF and MA assisted with writing the original draft and revising the manuscript. YG-M coordinated genetic testing, assisted in managing genetic data, and reviewed and revised the manuscript draft. JL provided statistical consultation and reviewed the statistical outputs. SD-F provided mentorship and reviewed and revised the manuscript draft. RNR led the parent study, provided mentorship, contributed to the study design and analytic plan, and reviewed and revised the manuscript draft.

## Competing Interests Statement

There are no competing interests related to this work to disclose.

## Funding Information

This study was funded by Columbia University’s and the New York State Psychiatric Institute’s HIV Center for Clinical and Behavioral Studies, a program funded by the National Institute of Mental Health (P30 MH043520, PI: Remien). This work was also supported by funding from a training grant from the National Institute of Mental Health (T32 MH019139, PI: Sandfort) and funding from the Eunice Kennedy Shriver National Institute of Child Health and Human Development (R21 HD098035, PI: Robbins).

## Author Declaration

This work was conducted under ethical approval from the IRB at the New York State Psychiatric Institute and received a Certification of Ethical Clearance from the Human Experimentation Committee at the Research Institute for Health Sciences, Chiang Mai University.

## Notes

**Funding and Conflicts of Interest.** This work was supported by funding from the National Institute of Health (P30 MH043520, PI: Remien; T32 MH019139, PI: Sandfort; R21 HD098035, PI: Robbins). Authors have no conflicts of interest related to this work to declare. Acknowledgments and declarations are included on the page immediately preceding references.

### Competing Interest Statement

The authors have declared no competing interest.

### Author Declarations

This study was approved by the New York State Psychiatric Institute's IRB and the Research Institute For Heath Sciences, Chiang Mai University's Human Experimentation Committee. Written informed consent was collected from participants >18 years. For participants <18 years, participant assent and written informed consent from a parent/guardian were collected.

